# Accurate Machine Learning Model for Human Embryo Morphokinetic Stage Detection

**DOI:** 10.1101/2024.11.04.24316714

**Authors:** H Misaghi, L Cree, N Knowlton

## Abstract

**Purpose:** The ability to detect, monitor, and precisely time the morphokinetic stages of human pre -implantation embryo development plays a critical role in assessing their viability and potential for successful implantation. Therefore, there is a need for accurate and accessible tools to analyse embryos. This work describes a highly accurate, machine learning model designed to predict 17 morphokinetic stages of pre-implantation human development, an improvement on existing models. This model provides a robust tool for researchers and clinicians, enabling the automation of morphokinetic stage prediction, standardising the process, and reducing subjectivity between clinics.

**Method:** A computer vision model was built on a publicly available dataset for embryo Morphokinetic stage detection. The dataset contained 273,438 labelled images based on Embryoscope/+© embryo images. The dataset was split 70/10/20 into training/validation/test sets. Two different deep learning architectures were trained and tested, one using EfficientNet-V2-Large and the other using EfficientNet-V2-Large with the addition of fertilisation time as input. A new postprocessing algorithm was developed to reduce noise in the predictions of the deep learning model and detect the exact time of each morphokinetic stage change.

**Results:** The proposed model reached an overall test accuracy of 87% across 17 morphokinetic stages on an independent test set.

**Conclusion:** The proposed model shows a 17% accuracy improvement, compared to the best models on the same dataset. Therefore, our model can accurately detect morphokinetic stages in static embryo images as well as detecting the exact timings of stage changes in a complete time-lapse video.

## Introduction

Time-Lapse Imaging (TLI) incubators were first introduced into human *in vitro* fertilisation (IVF) clinical laboratories around 2010[1]. Monitoring embryonic development through the capture of periodic images of each embryo, taken 5-20 minutes apart, holds promise, as the timings of specific developmental events have demonstrated associations with implantation potential, reviewed in [2]. Static images taken minutes apart can be assembled into a comprehensive video, chronicling the embryo’s *in vitro* progression. This offers a dynamic perspective on embryo development and growth.

TLI can be considered an advanced method for improving embryo quality assessment. It helps in selecting, deselecting and ranking embryos, potentially reducing the time to pregnancy during IVF treatments [3] [4] [5]. Numerous studies have identified an association between morphokinetic (MK) parameters and the likelihood of implantation [6], embryo ploidy [7], fetal-heartbeat [3], and live birth [4][5][8]. The volume of high-resolution imagery generated by TLI also offers opportunities for the application of Machine Learning (ML) and Artificial Intelligence (AI) techniques, including Deep Learning (DL), to improve embryo selection by increasing reproducibility and reducing human effort.

Several methodologies have been proposed to provide an accurate assessment of embryo viability, operating on the premise of accurately annotated timings of morphokinetic events[9] [10]. Rubio et al. conducted the first randomised control trial to determine the efficacy of a multivariable morphokinetic model on success rates. The authors found a significant increase in implantation and ongoing pregnancy rates and a significant decrease in early pregnancy loss for the cohort using TLI combined with multivariable models based on morphokinetic timings [11]. Shortly after, computational combinations of morphokinetic timings were integrated into a Known Implantation Determination Day 3 Score (KID D3 Score). This score uses a decision tree based on timings to get to and interval between MK stages including time to pronuclear fading (tPNf), two cells (t2), three cells (t3), and five cells (t5), to predict embryo viability [12]. Reliability of manual morphokinetic stage annotation is variable, with good agreement at specific time points including t2, t3 and t4 and less agreement at tPNa (time of pronuclei appearance) and t9+ [12] [13]. This variability, coupled with the labour-intensive nature of manual annotation, highlights the need for automated solutions, such as those enabled by ML and AI.

### Automatic Morphokinetic stage detection

Machine learning methodologies have been employed at various stages of embryo selection processes. Most focus directly on predicting the success rate of an embryo reaching the fetal heartbeat stage [14]. Some tools have been reported to surpass embryologists in accurately identifying viable embryos[15]. There are three main approaches to utilising TLI for embryo selection. The first approach is models working directly with the videos from TLI images, requiring no frame selection by the user [15] [16]. The second is methods that work on a single image where the user needs to select the specific frame from the TLI images for predictions [17][18]. The third is a hybrid approach where the timings are extracted manually and then fed into a ML model such as Kidscore [19] [9].

While approaches such as IDAScore V1 [15] and V2 [16] use time-lapse videos, others including ERICA [20], Life Whisperer [17], and Stork [18] rely on a single static image at the blastocyst stage of development. The pipeline for selecting the blastocyst image is not fully automated for ERICA [20], LIFE whisperer [17], and Stork [18], necessitating manual selection. However, automating the detection of morphokinetic events could streamline the entire process by identifying the most appropriate images and incorporating them into the pipeline. This hybrid approach involves extracting timings and feeding into a ML model such as Kidscore [19] [9].

Early methods of automatic morphokinetic stage detection relied on biological features to identify morphokinetic events [21]. For example, Feyeux et al. used grey-level analysis of microscopic images to predict morphokinetic stages [10]. In recent years, however, modern approaches have primarily utilised deep learning techniques [14], focusing on the supervised training of convolutional neural networks to facilitate the automatic annotation of morphokinetic events. Models employing ResNet-50, Long Short-Term Memory (LSTM), and ResNet-3D are now commonly adopted [3].

Although the models that predict morphokinetic events show similar levels of accuracy and overall performance, most are developed on proprietary datasets that are not publicly available. Two prominent examples are the commercial models for EmbryoScope [21] and Gerry Incubators [22]. Zabari et al. proposed a DL methodology that uses video frame-based initial predictions, which are further refined through monotonic regression. This approach aims to mitigate prediction noise by limiting predictions to either the current or an immediately subsequent developmental class [21].

Recently, Gomez *et al.* has introduced a large, annotated dataset, comprising 704 videos of developing embryos, featuring 337,000 images across 16 developmental stages. This dataset provides a critical resource for model benchmarking and development. In this current study multiple different DL models will be developed and trained on the Gomez et al. dataset [3], and evaluated. This novel, multimodal methodology aims to incorporate both embryo images and the time since fertilisation as inputs, to enhance the model’s performance.

## Material and Method

### Dataset

Time-lapse images of human embryos were obtained from the publicly available dataset (Gomez et al. [3]). The dataset contains 704 Embryoscope videos recorded at 7 focal planes and annotated for 16 morphokinetic events (Vitrolife ©). A single embryologist labelled all videos. To label the videos, the embryologist first identified the frame in which each event occurred and assigned the label to these frames. Then, all subsequent frames until the next morphokinetic event occurs were assigned the current label [3].Each video contains on average, 8 morphokinetic events. Of the total videos, 499 show viable embryos, with the remaining 205 showing non-viable embryos, capturing the myriad features that occur during embryonic development, including features such as levels of fragmentation, tri-pronuclear (3PN) fertilisation, and necrosis, etc. During the current study, only the central focal plane was used for labelling and only files with uncorrupted jpeg images were analysed, resulting in 273,438 images with labelled events.

To mitigate some of the subjectivity in identifying morphokinetic transitions, two images before and two images after the recorded transition time were removed from the training set. For the evaluation step, all images were included in the test set, with no exclusions around the transition points.

During the dataset quality review, it was noted that, as is standard practice, the embryos are removed for freezing or transferring on Day 5; however, the label still reflects the last event, i.e. expanded blastocyst. This mislabelling of the wells introduces ground truth errors in the dataset. To identify and relabel these images, an empty/non-empty model was developed and applied to all images (Appendix 1). The results were verified visually, and 9,734 images were labelled as empty wells.

Additionally, it was noted that there were only 41 hatched blastocyst images (Table 1). This small number of examples makes predicting this class difficult and subject to increased variability therefore it was not analysed in further sections.

**Table 1.** Updated Gomez dataset definitions and number of samples for each class in the train, test, and validation sets.

| Annotation | Description | Number in Training set | Number in Test set | Number in Validation set |
| --- | --- | --- | --- | --- |
| tPB2 | Polar body appearance | 5641 | 1737 | 880 |
| tPNa | Pronuclei appearance | 27762 | 8156 | 3839 |
| tPNf | Pronuclei disappearance | 4411 | 1279 | 643 |
| t2 | Cleavage stage 2 cells | 18968 | 5417 | 2825 |
| t3 | Cleavage stage 3 cells | 2825 | 958 | 570 |
| t4 | Cleavage stage 4 cells | 18483 | 5366 | 2815 |
| t5 | Cleavage stage 5 cells | 5009 | 1336 | 607 |
| t6 | Cleavage stage 6 cells | 5154 | 1657 | 758 |
| t7 | Cleavage stage 7 cells | 7094 | 1810 | 721 |
| t8 | Cleavage stage 8 cells | 20133 | 5276 | 3250 |
| t9+ | Cleavage stage more than 9 cells | 31241 | 9383 | 4579 |
| tM | Morula | 10402 | 3061 | 1608 |
| tSB | Start of Blastulation | 10206 | 2955 | 1709 |
| tB | Blastocyst | 4796 | 1096 | 556 |
| tEB | Expanded blastocyst | 11002 | 3928 | 1761 |
| tHB | Hatched blastocyst | 32 | 9 | 0 |
| Empty | Empty well | 7113 | 1847 | 774 |

Table 1 presents the dataset annotations, including the addition of the “empty” class, and the number of samples corresponding to each class.

### Deep learning models architecture

For Model 1, the backbone is an EfficientNet-V2-Large [23] fine-tuned to categorize 17 morphokinetic classes as outlined in Table 1. The input to the model was static greyscale JPG images of 380×380 resolution and the greyscale values were copied 3 times into the R,G,B channels. Weights were initialised from a network pre-trained on ImageNet[23]. Figure 1(a) illustrates the structure of this network.

**Figure 1.**
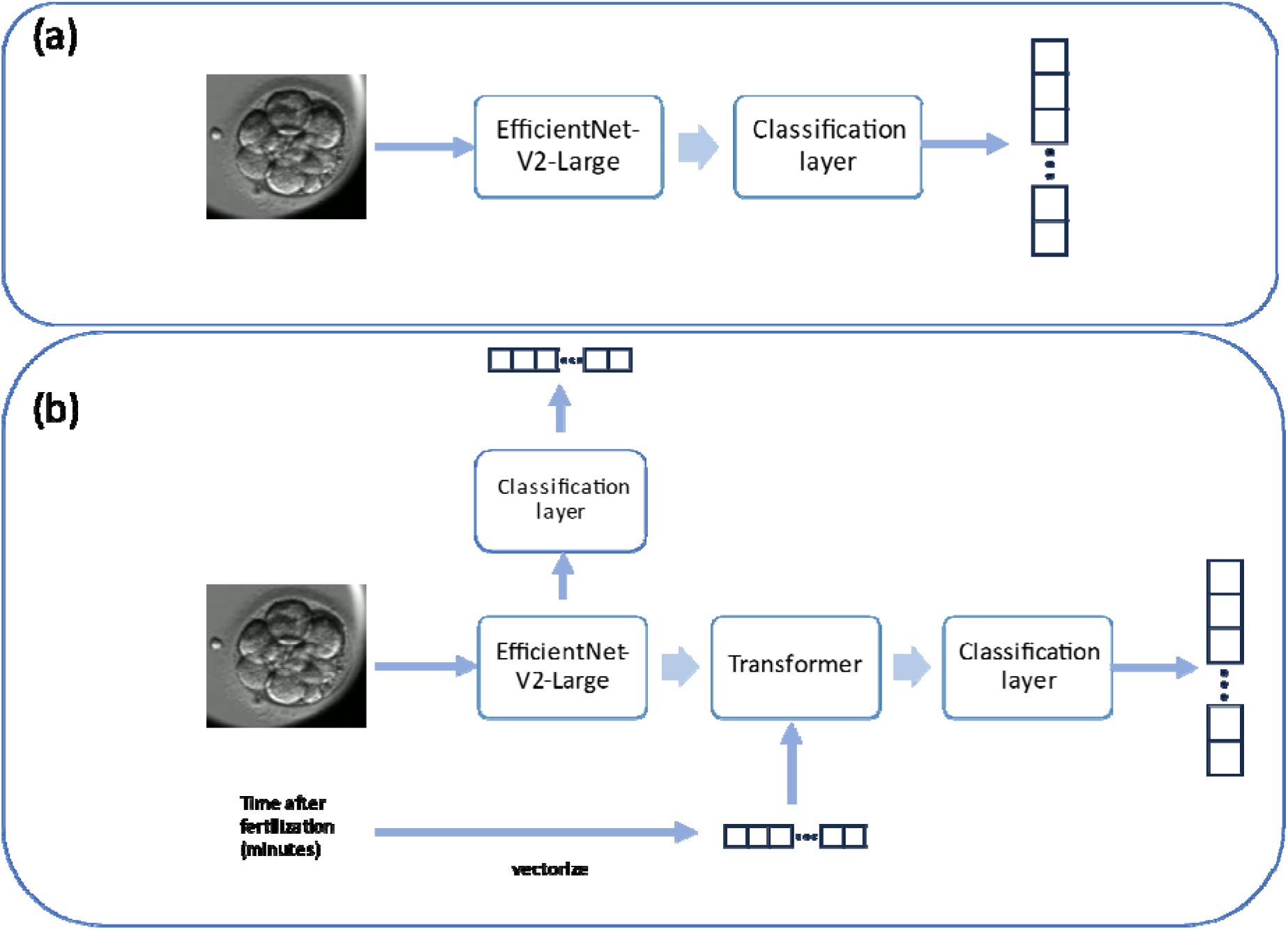
Architecture of the proposed models (a) Model 1, utilises EfficientNet-V2-Large as backbone for feature extraction and it has a fully connected layer as the classifier head. (b) Model 2, has EfficientNet-V2-Large as a feature extractor, this model has two classifier heads one after the feature extraction step and one after the transformer. The transformer in model 2 fuses the time after fertilization and extracted features from the EfficientNet-V2-Large. The two head structure of this network ensures the proper gradient flow to backbone.

Model 2 also uses an EfficientNet-V2-Large as the backbone for processing images, followed by a transformer [24]. The transformer has only one encoder layer with a hidden size of 512 with 4 self-attention heads. The second input is the number of minutes that have elapsed since the time of fertilisation. As transformers are designed to work on sequences of vectors, the time since fertilisation is converted into a binary vector representing two-hour windows of time. This approach allows the self-attention mechanism to interact with the image features extracted earlier in the pipeline. A maximum incubation time of seven days was assumed. Time in hours was then encoded into a two-hour window using a one-hot vector or length, resulting in a vector length of 84.

The time vector is calculated as follows:

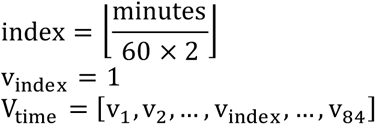

Where all v elements are 0 except for v_index_. For example, if the time to reach a stage is 150 minutes, the index will be calculated as 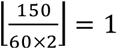, and [0, 1, 0, 0, …, 0]. This time vector will be the same for 160 minutes, but for 240 minutes the index will be 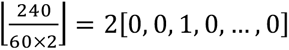.

The time vector and the image features generated by the backbone are passed to the transformer layer (Figure 1 (b)). A final Multilayer Perceptron (MLP) is then used to make class predictions for each image. By design, this model has two classification heads, one immediately after the backbone and one after the transformer layer. During training, two cross-entropy losses are summed. This ensures proper gradient flow to the backbone layers and prevents the model from overfitting only on the time vector inputs.

Model 3 meanwhile has the same architecture as Model 1, but was trained on the original Gomez dataset that did not include the reclassified ‘empty’ well labels. Model 4 was trained with a Resnet-50 backbone to replicate the previous work by Gomez et al. [3], and to serve as a baseline model for comparison.

All models were trained using the Adam optimiser, as provided in the PyTorch 2.0.0 library [25], with an initial learning rate of 0.001 and a Cross-entropy loss function. The learning rate was dynamically reduced upon observing a plateau decrease in the validation set performance. The networks underwent training for 50 epochs on the training dataset, which was found to be sufficient as validation loss stabilised with no significant improvements were observed beyond this point. Models were subsequently evaluated on the test set, which remained unseen during the training phase.

The issue of class imbalance represents a significant challenge within this dataset, as illustrated in Table 1. Such imbalance can induce bias in the neural network towards classes with a higher proportion of samples. To mitigate this issue, the inverse class frequency method was used. This technique enables the model to generate appropriate gradient adjustments for the classes that are represented by fewer samples.

Embryo images often exhibit significant variations in brightness, with some regions appearing overly bright and others notably dark. Therefore, proper image normalization is crucial. The image augmentation techniques employed during training included rotating and shifting the image with a 30% probability, flipping the image with a 50% probability, and applying noise or blur with a 50% probability, followed by a final normalisation step to ensure consistent image quality across the dataset. The augmentation step was conducted using the Albumentations library in Python [26]. The normalisation step was performed using image contrast enhancement (CLAHE, Contrast Limited Adaptive Histogram Equalisation) and the normalise function, with average pixel values across RGB channels set as (0.485, 0.456, 0.406) and the standard deviation as (0.229, 0.224, 0.225) to align with the ImageNet weights.

### Postprocessing algorithm

The morphokinetic annotation of videos critically depends on the consistency of predictions across all video frames. Given the dynamic nature of embryo development, there are instances when the classification of a frame is ambiguous, resulting in “noisy” predictions. To accurately assess an embryo’s viability using methods like KidScore, it is crucial to accurately identify the exact timepoint when the morphokinetic stage of the embryo changes[12]. To address this a heuristic method was developed. This method is utilised on the predictions made by networks for each frame of the embryo’s timelapse video. The algorithm accounts for transitions to more advanced morphokinetic stages and instances of reverse cleavage. It therefore identifies a trend within each time interval and corrects mispredictions that may disrupt this trend. Mispredictions often occur due to embryo movement or the movement of cells within it, for example, where some cells may be temporarily obscured in the image. This would render morphokinetic stage changes inaccurate.

The algorithm works in two stages: First, it amends the predictions by substituting low-confidence predictions with the latest morphokinetic state prediction with greater than 80% confidence. Second, it detects changes by comparing the predicted class with the prediction of the preceding images.

The heuristic is defined as follows:

Let P_1_ represent the value of the most probable predicted class at the index i, changes are defined as Δ(Equation 1):

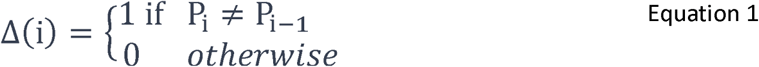

By summing the values of changes within a sequence, various consecutive groups can be delineated as follows (Equation 2):

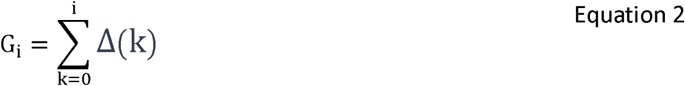

The values of G_I_ can be used for grouping, indicating that all images with identical G_I_ values form a continuous group and share prediction values without interruption. By comparing the length of each continuous group with its neighbouring groups, it can be determined whether the current group should be considered noise or retained as a valid morphokinetic stage change. The algorithm is outlined as follows:

- Let C = {c1, c2, …, cn} represent the set of unique predicted classes in the video.
- For each class c in C, create a subset Dc of all the predictions D such that all elements in Dc have prediction class equal to c or each subset Dc, identify unique group values as Gc = {g1, g, …, gm}.
- Calculate the length of each consecutive group g in Gc, denoted as Lg, where Lg is the number of elements in Dc that belong to group g.
- Determine the minimum and maximum values within G_c_, denoted as g_min_ and g_max_, respectively.
- For each potential group identifier i_g_ in the range [g_min_, g_max_], identify if i_g_ is an interruptive group by checking if ig ∉ Gc. For each interruptive group, calculate its length L_ig_.
- All interruptive groups are discarded and only the main groups are kept as final labels.

Figure 2 shows an example of the model’s predictions in blue, highlighting instability at certain points throughout the video, particularly near morphokinetic transition times. To address this, a post-processing algorithm has been developed to refine these noisy predictions. The result is a stable line shown in orange, ensuring that the morphokinetic stage detection outputs are reliable for further use.

**Figure 2.**
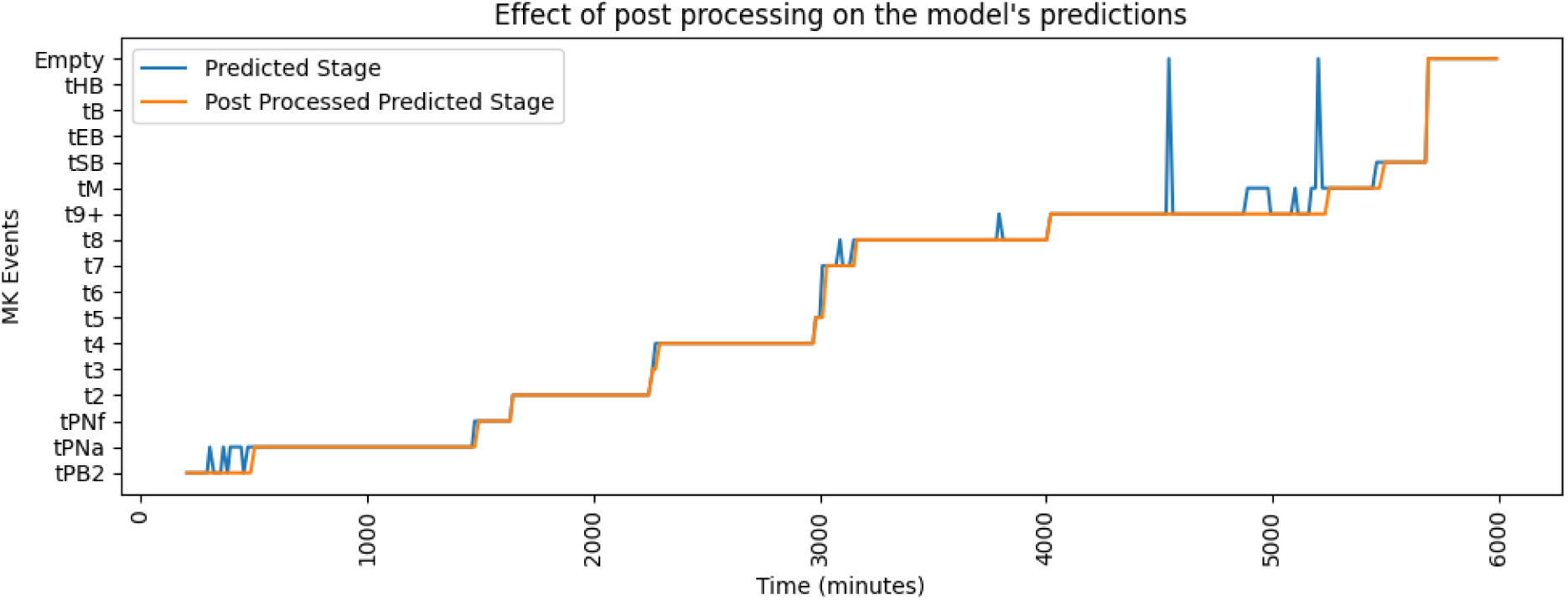
An example of the effect of the postprocessing algorithm on the predictions generated using Model 2. The model predictions are uncertain at times throughout the video, specifically near a morphokinetic stage change (blue). The postprocessing algorithm ensures a clean set of predictions by assessing and refining noisy predictions (orange).

## Results

After 50 epochs of training, Model 1 showed an accuracy of 93% while Model 2 showed an accuracy of 95%; Models 3 and 4 had an accuracy of 93%, and 96%, respectively, on the training set.

Next, each model’s network was evaluated on a subset of data which remained unseen by the models during the training phase (test set). To properly assess the models and postprocessing algorithm the evaluation steps are separated, and results are presented in two sections; The first is single image processing, where the results are based on deep learning models of single images. The second is postprocessing, where results are presented after applying the model on a video created by static time lapse imaging. The postprocessing algorithm is used to extract the exact time of morphokinetic stage changes.

### Single Image Processing

During the testing step standard classification metrics were calculated for each model. All the images in the test dataset were processed and the outputs compared against ground truth. A confusion matrix, accuracy, F1-score, precision, and recall were calculated for each model. Figure 3 presents the confusion matrix of Model 2, the highest performing model, when it was applied to the test dataset. Certain stages, such as t2 and t4, were easier to identify whereas other stages, including tB and t5, are more challenging for the model to accurately classify. Misclassifications most commonly occur between consecutive stages, such as when the images of class t5 are misclassified as either t4 or t6. However, it is highly unlikely for the model to confuse an image with a class significantly ahead or behind. For example, an image labeled as t5 has a less than a 4% chance of being classified as anything other than t4 or t6.

**Figure 3.**
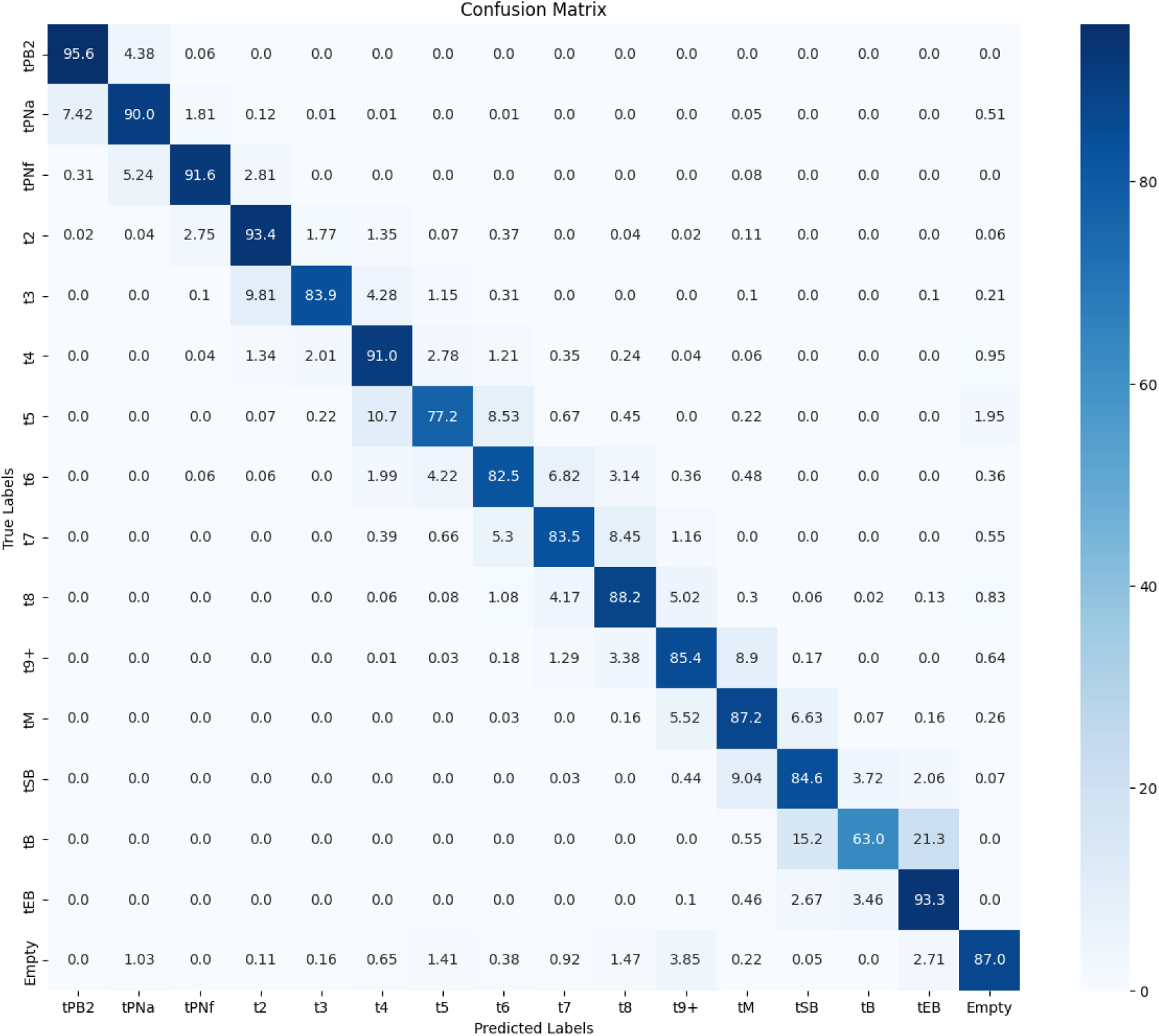
A confusion matrix for Model 2 on the test set in percentages. The blue gradient bar on the right of the matrix indicates the percentage graphically with 0% - white and 100% - dark blue. Values of 0.0 indicate that no misclassification occurred, and therefore the model prediction was accurate, down the lower right diagonal where values would be 100%.

Standard classification metrics for the two highest performing models are shown in Table 2. Both models show high accuracy levels, with Model 2 demonstrating a slightly superior performance on all the metrics.

**Table 2.** Model performance of the top two models across a range of performance metrics. Note accuracy here is calculated across all 17 classes.

| Model | Precision | Recall | F1-Score | Accuracy |
| --- | --- | --- | --- | --- |
| Model 1 | 0.883 | 0.871 | 0.873 | 0.871 |
| Model 2 | 0.886 | 0.879 | 0.881 | 0.879 |

Comparing the current models with those trained using the Gomez et al. model, Models 1 and 2 demonstrate an approximate 17% improvement in accuracy (Table 3). Models 1 and 2, which contained the empty well classifier had the best performance, whereas Models 3 and 4, were less accurate. Model 4, which was trained to represent the Gomez work, is performing only slightly better than the resnet-50 that was reported by Gomez et al [3].

**Table 3.** Accuracy comparisons between all models and those reported in Gomez et al [3]. All methods were trained on the same publicly available dataset[3]. Models prefixed with Gomez is reported performance on this dataset for comparison purposes. Models 1 and 2 were developed to improve discriminatory accuracy. Models 3 and 4 are comparable to the Gomez models directly, as they don’t include our updated class of “empty well”.

| Method | Test set Accuracy |
| --- | --- |
| Gomez et al. ResNet | 0.663 |
| Gomez et al. ResNet-LSTM | 0.685 |
| Gomez et al. Resnet-3D | 0.705 |
| Model 1 (Efficient net) | 0.871 |
| Model 2 (Efficient-net-transformer modality) | 0.879 |
| Model 3 (Efficient net, without empty well labels) | 0.739 |
| Model 4 (Resnet-50, without empty well labels) | 0.689 |

To visually assess the root causes of misclassification of the models, a random sample of images that were mis-predicted by Model 2 for the classes that had highest error rate are shown Supplementary Figure 1 to Supplementary Figure 4.

### Postprocessing

After applying the postprocessing method the exact predicted times for each morphokinetic event was extracted. In the analysis only the resulting morphokinetic state is considered, for example, changes from t2 to t4, and t3 to t4 are considered as transition to t4.

While the model results were accurate, a plus or minus one developmental stage classifications was observed in a sequential set of images, as detailed above (Figure 3). This is expected due to embryo movement during culture, thus leading to obscurement of cells and features. To combat this, a heuristic post-processing was implemented.

Upon applying the model and subsequent postprocessing algorithm to all videos in the test dataset, the timings for morphokinetic stage changes were extracted. Error is calculated as (*time_prediction_* − *time_ground truth_*). The average, standard deviation, and percentiles (25%, 50%, 75%, 80%, 85%, 90%, 95%, and 99%) of the errors for each class are presented in Table 4. The results demonstrate that errors are small for the majority of cases, with large errors occurring in less than 5%. The largest errors were observed in the tB class.

**Table 4:** Quantile analysis of timing prediction errors for each morphokinetic class. Count shows the number of events of the specified class. Mean is the average error and std is standard deviation (in hours) for the class. Percentiles (25%, 50%, 75%, 80%, 85%, 95%, and 99%) are also reported for each class.

| event | tPNa | tPNf | t2 | t3 | t4 | t5 | t6 | t7 | t8 | t9+ | tM | tSB | tB | tEB |
| --- | --- | --- | --- | --- | --- | --- | --- | --- | --- | --- | --- | --- | --- | --- |
| count | 140 | 121 | 136 | 51 | 114 | 66 | 68 | 70 | 112 | 144 | 109 | 95 | 27 | 60 |
| mean | 0.54 | -0.01 | 0.26 | 0.23 | -0.74 | -0.87 | -0.45 | -0.07 | -0.78 | -2.94 | -1.03 | 0.61 | 1.61 | -1.51 |
| Std | 3.43 | 0.93 | 1.40 | 2.01 | 5.22 | 3.31 | 1.65 | 1.36 | 3.86 | 7.84 | 3.45 | 2.33 | 6.10 | 5.89 |
| 25% | 0.00 | -0.30 | 0.00 | 0.00 | -0.30 | -0.45 | -0.57 | -0.30 | -0.85 | -3.08 | -3.00 | -0.30 | -0.15 | -1.43 |
| 50% | 1.00 | 0.00 | 0.20 | 0.00 | 0.00 | 0.00 | 0.00 | 0.00 | 0.00 | 0.00 | -0.80 | 0.20 | 1.00 | -0.40 |
| 75% | 2.00 | 0.20 | 0.30 | 0.30 | 0.30 | 0.20 | 0.30 | 0.60 | 0.70 | 0.85 | 0.20 | 1.10 | 2.60 | 0.70 |
| 80% | 2.04 | 0.20 | 0.50 | 0.30 | 0.50 | 0.30 | 0.46 | 0.72 | 0.70 | 1.20 | 0.74 | 1.82 | 3.10 | 1.02 |
| 85% | 2.50 | 0.30 | 0.50 | 0.70 | 0.70 | 0.53 | 0.60 | 1.00 | 1.07 | 1.76 | 1.28 | 3.00 | 7.15 | 1.41 |
| 90% | 2.61 | 0.60 | 0.80 | 1.00 | 1.20 | 0.85 | 1.00 | 1.20 | 1.50 | 2.67 | 1.74 | 3.88 | 9.10 | 2.05 |
| 95% | 3.21 | 1.70 | 1.78 | 1.85 | 2.24 | 1.43 | 1.63 | 1.50 | 2.84 | 4.29 | 3.46 | 4.56 | 11.40 | 4.03 |
| 99% | 10.79 | 2.44 | 3.71 | 7.05 | 9.51 | 4.84 | 1.80 | 2.00 | 7.37 | 8.30 | 6.10 | 6.17 | 12.89 | 8.20 |

Time extraction showed varying degrees of accuracy across different classes. Some classes, such as tPNf, demonstrated exceptional accuracy with only −0.01 hours average error, while others like t9+ showed more significant discrepancies with −2.94 hour average error. It is important to note that images were captured 20 minutes apart, meaning only a single frame misclassification contributes 0.33 hours of error.

Due to the significant impact of labelling errors on the accuracy of stage change time extraction, it was important to conduct an in-depth assessment of label subjectivity, to identify the primary causes of model misclassification. Supplementary Figure 5 and 6 provide examples of the largest errors that were observed. These figures demonstrate examples of images (frames) from classes which had the greatest time detection discrepancies. These visual examples help highlight the inconsistencies in labelling and the challenges these inconsistencies pose for model accuracy.

## Discussion

Morphokinetic timings have been linked to predicting the development of embryos to the blastocyst stage, [19] and their implantation potential [8]. However, current models largely rely on subjective manual annotations, which may limit their accuracy and reproducibility. The association between morphokinetic parameters and developmental outcomes could shift if more objective and standardised timings were used [8] [27]. Manual annotation is inherently subjective, and often only a limited set of annotations are performed, potentially omitting critical data. To advance the field of embryo morphokinetic research, it is crucial to develop models that are tested and validated on open-source datasets. While commercially available morphokinetic models like Fairtility [21] and the Gerry TLI system [22] exist, they are proprietary, and their performance has not been assessed on publicly available datasets, limiting their transparency and comparability.

Table 3 provides a comparative analysis between the models introduced in this study and those developed by Gomez et al. [3], highlighting the improved performance accuracy of Model 2, with 87.9 % accuracy on single frames. Since the same publicly available dataset was used for training and testing as in Gomez et al. [3], a direct comparison was feasible. However, a direct comparison with the study by Zabari et al. [21] may not be entirely valid, the accuracy of Model 2 (87.9%) is broadly comparable with the levels reported by Zabari et al. [21] (94%). However, our model has several advantages. First, it can detect reverse cleavage or any reduction in morphokinetic stage. Second, it can identify difficult stages such as the 9 cell, and intra-blastocyst stages (early, expanding, etc). This decreases our accuracy overall but allows the identification of potentially meaningful associations with embryos of poorer quality. It is also important to note that without evaluating the models on the same dataset, the performance metrics must be interpreted as a general indication only. Direct comparison is not meaningful as their study contained fewer morphokinetic classes than the dataset used in this study and consisted of 20,253 labelled embryos versus the 704 embryos in the publicly available dataset.

Although the dataset used to test the model from the Zabari et al. study is not publicly available, the current Gomez class definitions were adjusted to be aligned with the Zabari study, allowing the accuracy and morphokinetic timing detections to be compared. Using the Zabari et al. defined labels, the accuracy of our Model 2 is 91 % on the test set. This illustrates how these models are quickly converging to the Zabari model with 100 times less data. As described in Table 3, Model 3 has the same architecture and training settings as Model 1. While the model achieved 73.9% accuracy on the test set of the original Gomez dataset, by adding the empty well class, which is only 3.5% of the images in the dataset, the accuracy increased to 87%. This shows how mislabelling can affect the whole network’s learning process.

Misclassifications are clustered around stages that are likely to cause confusion with human annotators examining a single focal plane image. Typically, these areas tPNa, t5, t7, tM, tEB, and tHB. The inaccuracy of the network on these classes is higher and disagrees with the ground truth labels; nevertheless, there is more consistency in the model classification than amongst human labellers [28]. For example, the difference between an expanded blastocyst (tEB) and blastocyst (tB) is more subjective than the difference between a pronuclei fading (tPNf) and a two-cell (t2) embryo. Accurate labelling is particularly critical for tPNa and tPNf phases because although they show small visual differences, they represent different biological processes and mislabelling leads to model confusion. Examples for such mislabelling in the dataset is demonstrated in Supplementary Figure 5 (tPNa stage) and Supplementary Figure 6 (expanded blastocyst) Despite these “errors” most images in the dataset do have the correct labels, showing that the network has learned the classes correctly and is able to generalise well on the test set.

The postprocessing algorithm proposed here does not limit the predictions of the embryo stages to be monotonically increasing as implemented in both Gomez et al. [3], and Zabari. [21] Annotations provided by our model can therefore be used to detect how often reversions such as reverse cleavage occur and the effects this might have on embryo viability [29].

The difference between the performance of Model 1 and Model 2 is within a margin of error, as the difference in accuracy is only 0.6 %. This suggests that features existing in the images are sufficient for morphokinetic stage detection and adding the data about the post-fertilisation time does not improve the results from the model. Given the strong relationship between the time from fertilisation and developmental stage, it is unclear why this does not improve the results, but this may be due to the 100s of embryos in the dataset that fail to develop into blastocysts, reducing the predictive ability of time. This lends support to the potential for Model 1 to generalise across clinics with different media and embryo conditions, factors which that may affect the timing of developmental events.

The precise timing of MK stages is of interest. By applying models and post-processing algorithms, it was possible to extract these timings accurately (< 1hr). Current commercial software, which uses a range of 150-400 min, suggests that this time frame is indeed acceptable in current lab practice [22]. More work on the Gomez dataset could help improve the annotations further, enhancing morphokinetic determinations across the field. For example, new annotations introduced here included wells that were too dark to see the embryo, and those wells without embryos, classified as “empty”. Using the Gomez dataset as an objective benchmark for commercial morphokinetic software would provide a robust, repeatable metric to assess this class of model. Additionally, it would provide insight into which features, stages, or images are most difficult to predict on a model-by-model basis.

There are several limitations to the current approach. First, the model relies on human-annotated ground truth labels which contain inherent subjectivity (Supplementary Figure 5 and Supplementary Figure 6), while the labels were cleaned, there is likely not a consensus embryo staging for each image, especially during stage transitions, i.e. t4-t5. Our mitigation attempt was based upon removing two images before and after the transition during training but predicting these transitions during evaluation on the test set. Second, our dataset is limited to images from a single device type, Embryoscope+, and current model might not be able to generalise well on the images captured by other time-lapse incubators.

## Conclusion

This study introduces a highly accurate machine learning model for detecting the morphokinetic stages of human embryos, achieving up to 87.9% accuracy and significantly advancing in vitro fertilization (IVF) technology. The inclusion of a novel post-processing algorithm, which is not constrained by monotonicity, allows for the detection of reverse cleavage events, providing a more nuanced understanding of embryo development. By automating and enhancing the annotation of morphokinetic stages—traditionally a subjective and labour-intensive process—the model not only improves dataset quality but also delivers more consistent and reproducible results than human annotators. This model performs 17% better compared to previous methodologies on the Gomez et al. dataset, potentially streamlining embryo selection in clinical practice.

The timing errors for most morphokinetic stages fall within a clinically acceptable range of 1 hour, demonstrating the model’s practical utility. Furthermore, by detecting complex developmental patterns such as reverse cleavage, our approach may reveal new biomarkers of embryo quality that were previously overlooked.

Looking forward, this technology could reduce the subjectivity in embryo assessment, potentially increasing IVF success rates while decreasing the time to pregnancy and the emotional and financial burden on patients. As research like this continues to develop, it promises to standardize embryo evaluation across clinics worldwide, advancing both clinical outcomes and our fundamental understanding of early human development.

## Data Availability

Data is available upon reasonable request to the authors.

## Supplementary Figures

**Supplementary Figure 1.**
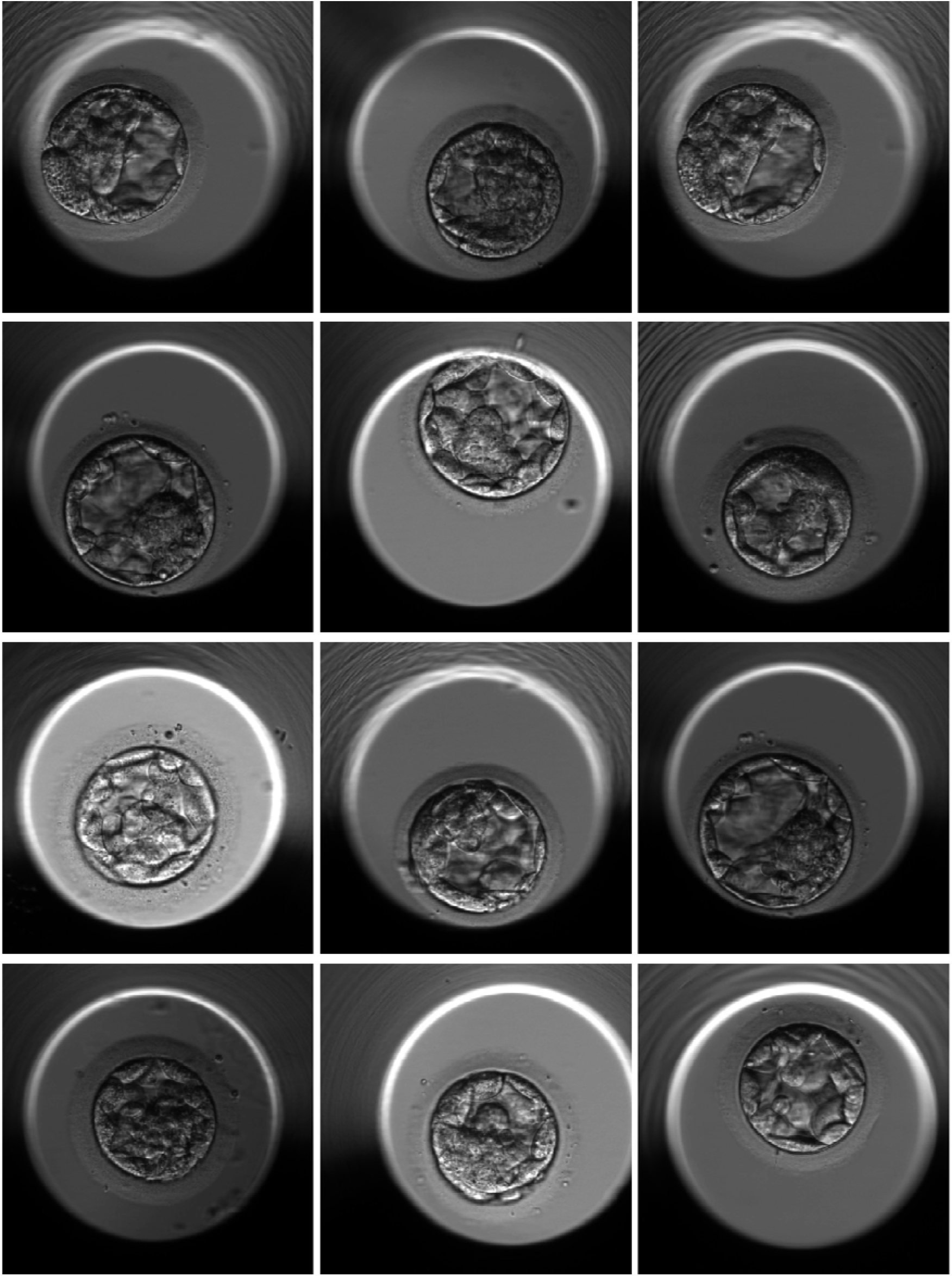
A random sample of Images was predicted as tEB by the network but was labelled as tB in the dataset. The misprediction between tEB and tB is among the worst classes for model 2, with 12% of the tB labelled images in the test set predicted as tEB.

**Supplementary Figure 2.**
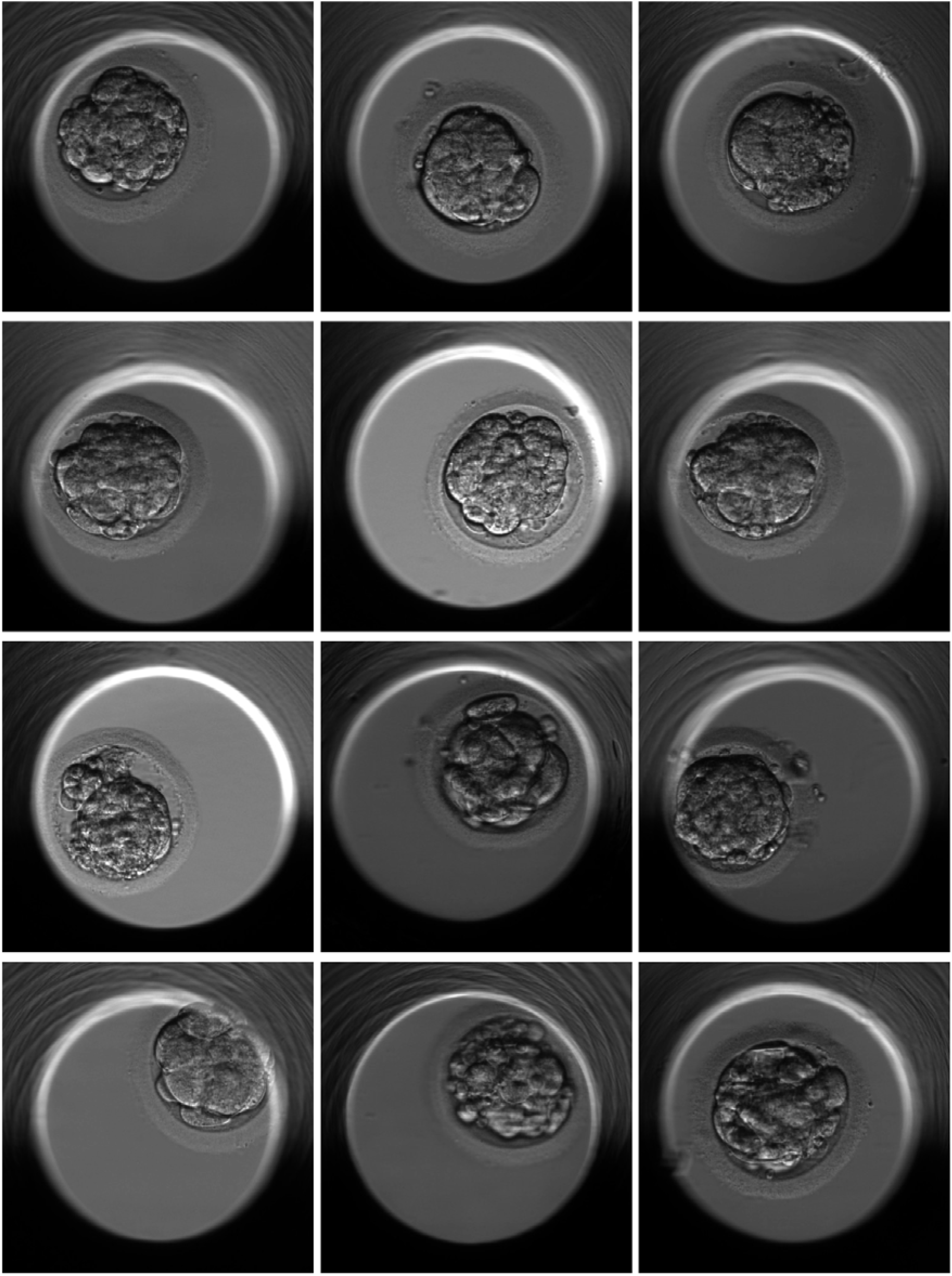
A random sample of Images that was predicted as tM by the network but was labelled as tSB in the dataset. The misprediction between tM and tSB is among the worst classes for model 2, with 12% of the tSB labelled images in the test set predicted as tEB.

**Supplementary Figure 3.**
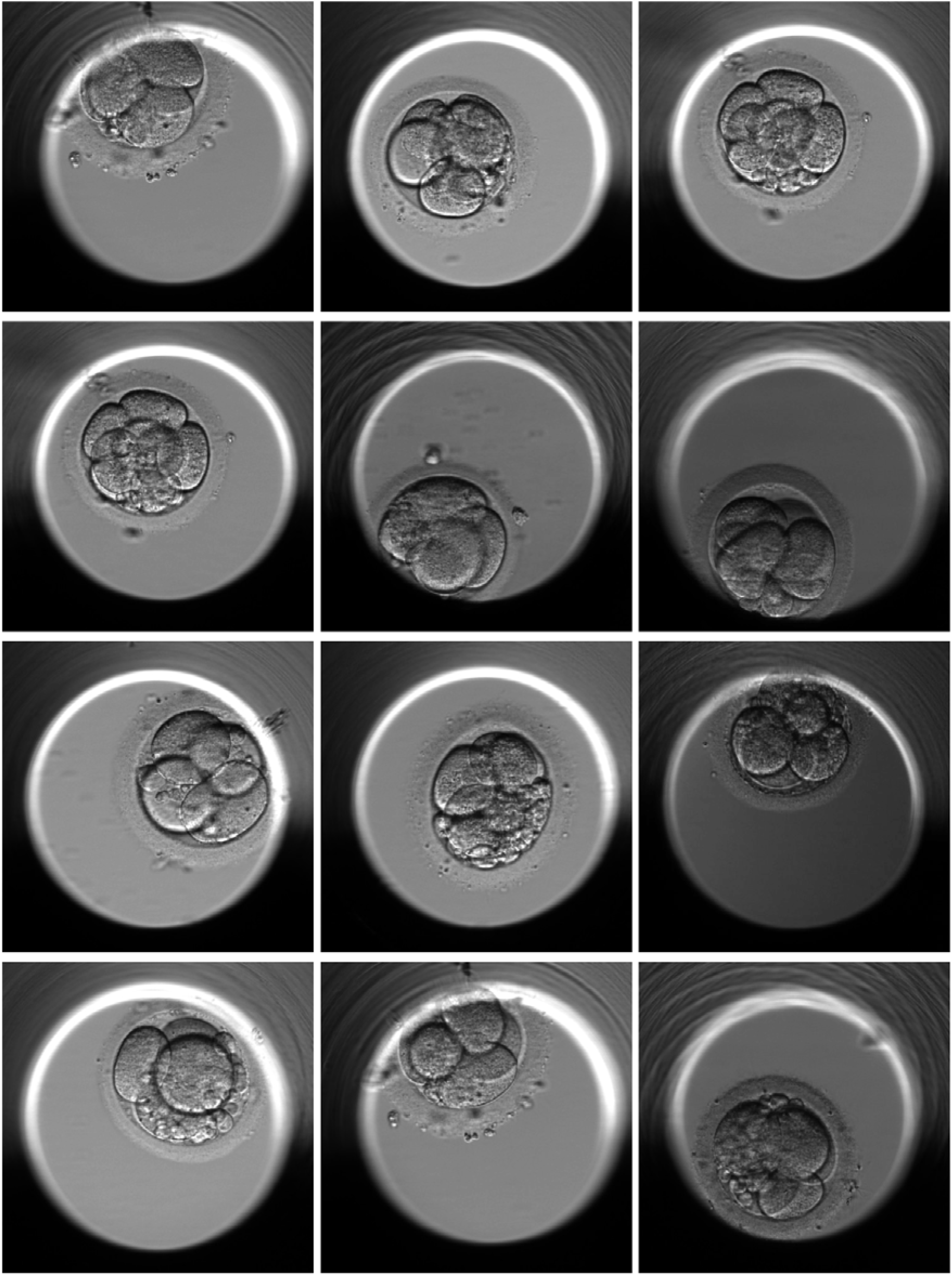
A random sample of Images that was predicted as t4 by the network but was labelled as t5 in the dataset. The misprediction between t4 and t5 is among the worst classes for model 2, with 16% of the t5 labelled images in the test set being predicted as t4.

**Supplementary Figure 4.**
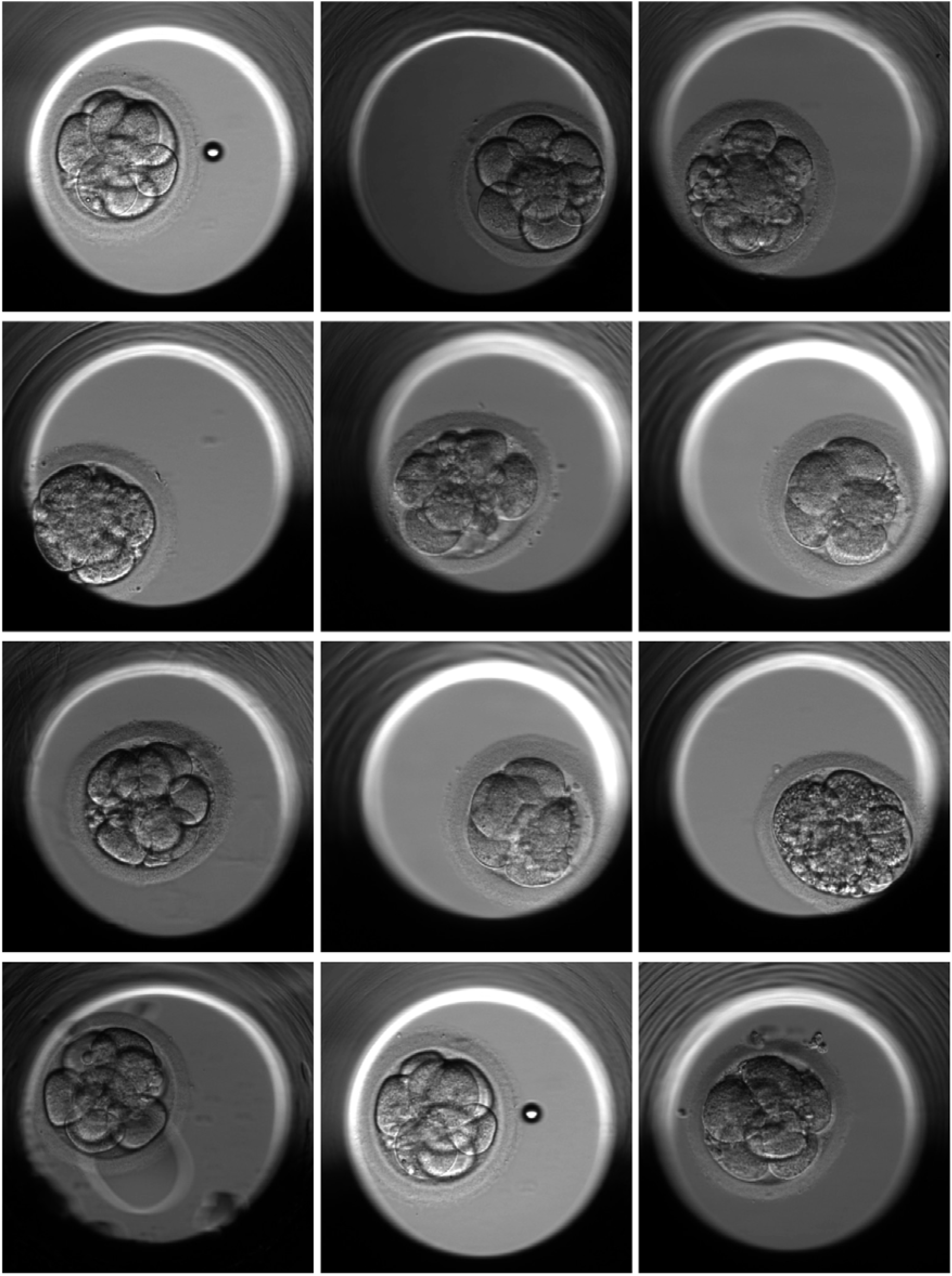
A random sample of Images that was predicted as t8 by the network but was labelled as t7 in the dataset. The misprediction between t8 and t7 is among the worst classes for model 2, with 13% of the t7 labelled images in the test set being predicted as t8.

**Supplementary Figure 5.**
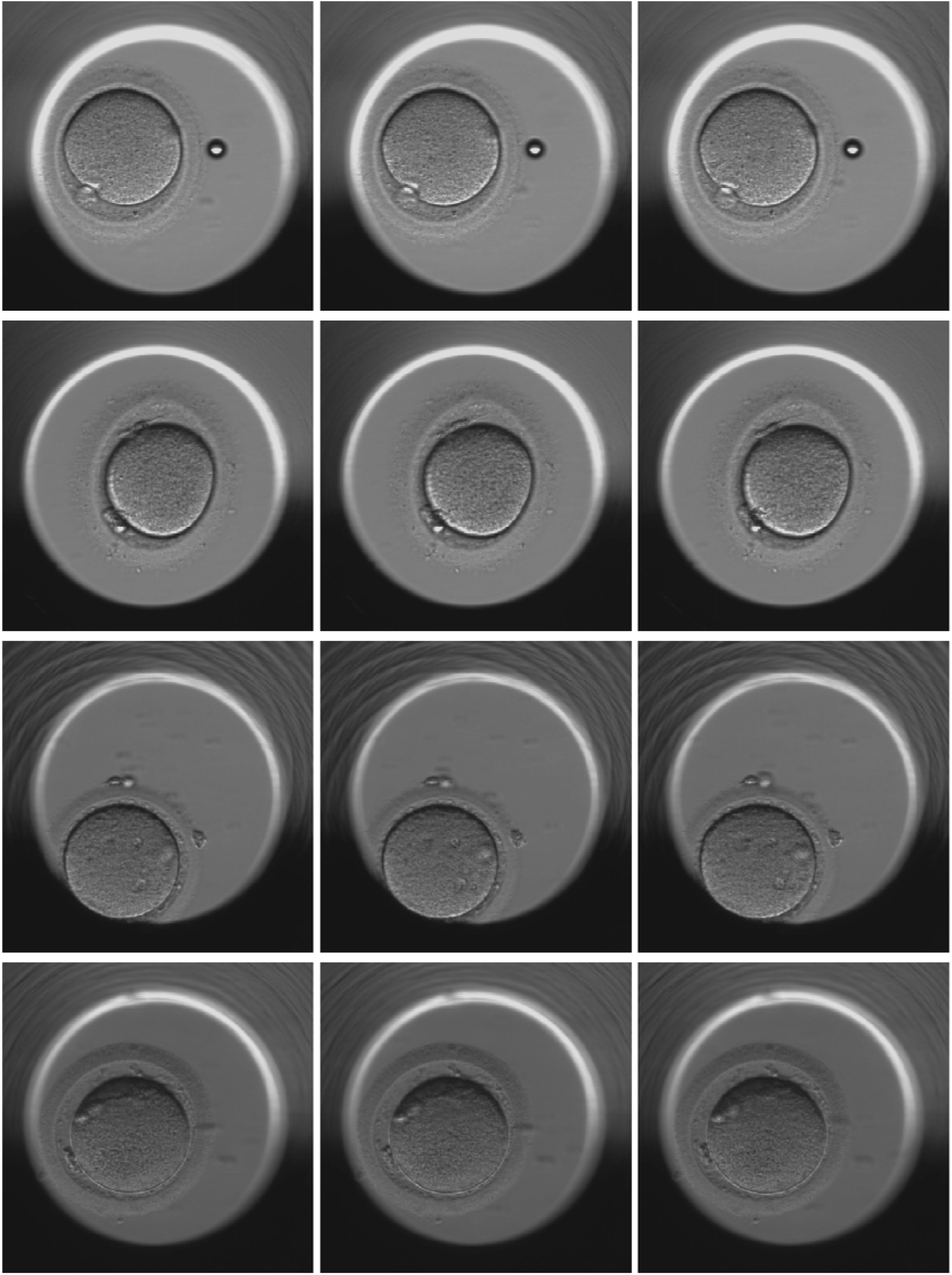
Examples from the dataset for embryos classified as tPNa. Each row corresponds to one embryo, and the middle image in each row corresponds to the exact frame reported in the dataset for the morphokinetic event changes. the image on the left is one frame before, and the image on the right is one frame after the labelled change. For this specific morphokinetic event, the pronuclei should be visible in the middle frame and after that.

**Supplementary Figure 6.**
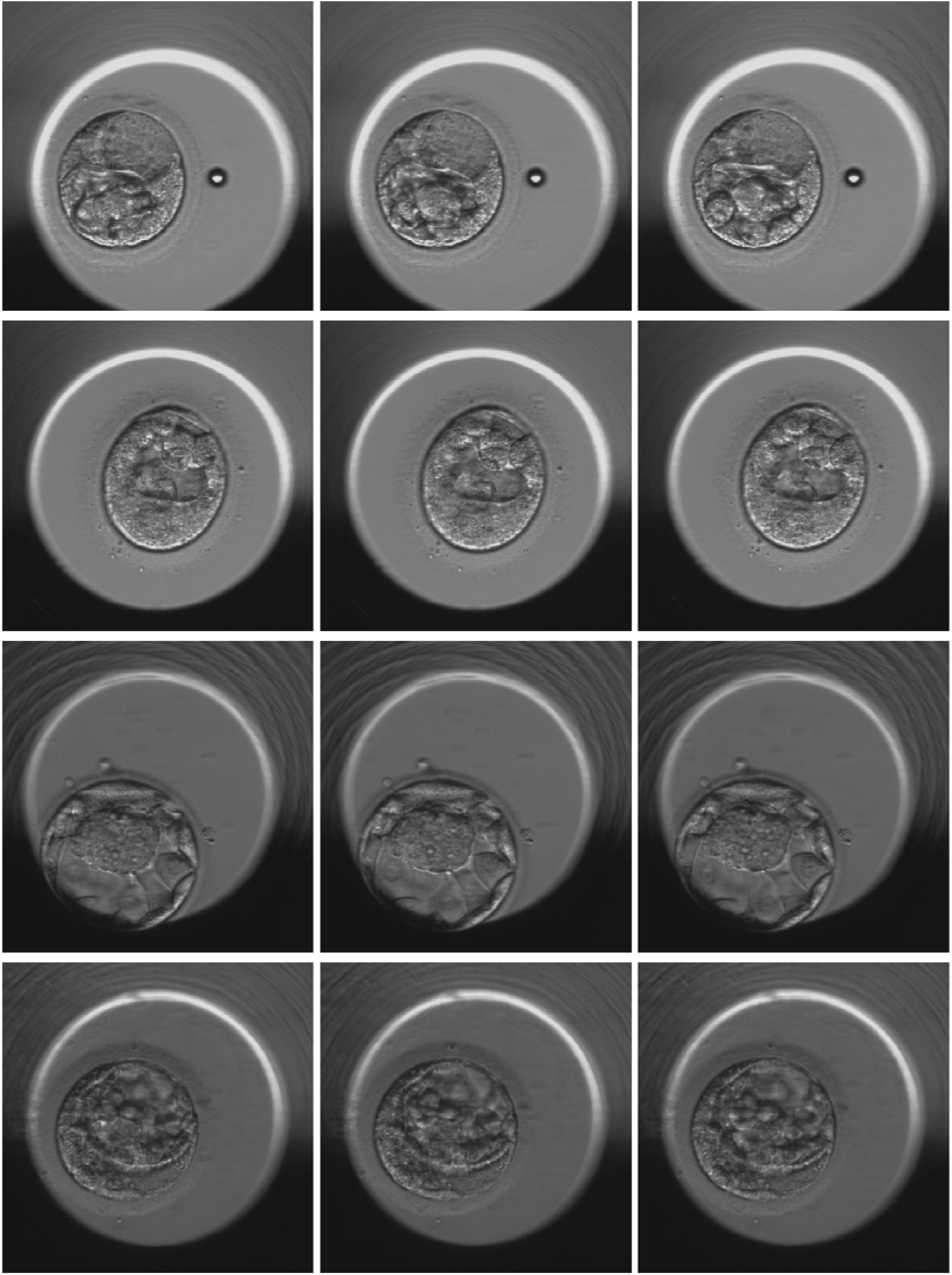
Examples from the dataset for class tEB. Each row corresponds to one embryo, and the middle image in each row corresponds to the exact frame reported in the dataset for the morphokinetic event changes. the image on the left is one frame before, and the image on the right is one frame after the labelled change. This shows the subjectivity in the classes between tB and tEB. It is unclear what the criteria for the difference between the classes is.

## Appendix 1

The model was trained on a dataset with 66,634 labelled images, the images were captured using Embryoscope device, this is the same type of device that was used in Gomez et al. dataset. This dataset was labelled with different set of label definitions in comparison to Gomez et al. dataset. Table 5 Shows the distribution of data and classes. This dataset includes labels for empty wells.

**Table 5.** Distribution of data in the second dataset.

| Classes | Description | Number of samples |
| --- | --- | --- |
| 2PN | Visible pronuclei | 6516 |
| Syngamy | Syngamy | 5950 |
| 2 Cell | Cleavage stage 2 cells | 7992 |
| 3 Cell | Cleavage stage 3 cells | 1761 |
| 4 Cell | Cleavage stage 4 cells | 5958 |
| 5 Cell | Cleavage stage 5 cells | 2340 |
| 6-7 Cell | Cleavage stage 6 or 7 cells | 4254 |
| 8 Cell | Cleavage stage 8 cells | 5296 |
| 9+ Cell | Cleavage stage 9 or more cells | 4577 |
| Compacted | compacted | 4023 |
| Blastocyst12 | Start of blasting and blastocyst | 6077 |
| Blast35 | Expanded blastocyst and hatched blastocyst | 3955 |
| Empty | Empty well | 7935 |

A classification network employing ResNet-50 [30] as the backbone was used to extract the features from images, then the classification is done using fully connected layers. This network used the same training methods and loss function as Models 1-3. For training this network the dataset was split to train and test subsets, 80 % were used for training and 20 % for testing.

**Figure 4.**
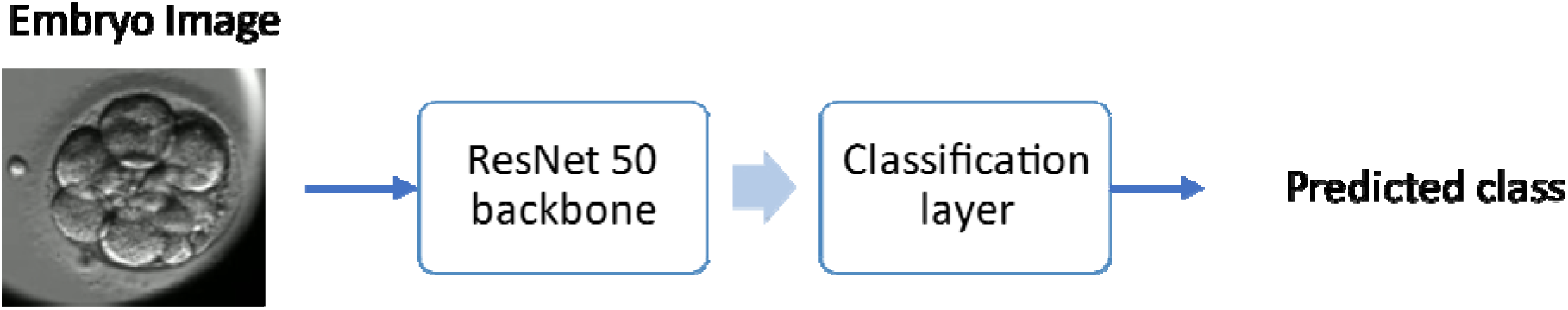
Model 1 Trained on Resnet-50

This model was able to predict the empty wells on the test dataset with 100% accuracy.

